# Precision Anti-Inflammatory Therapy in Atherosclerosis: A Systematic Review and Meta-Analysis of Colchicine Timing and Clinical Outcomes in Patients with Atherosclerotic Cardiovascular Disease

**DOI:** 10.64898/2026.03.25.26348968

**Authors:** Piyush Puri, Himanshi Yadav, Meet Kachhadia

## Abstract

**Background:** Despite optimal lipid-lowering and antithrombotic therapy, substantial residual cardiovascular risk persists in established atherosclerotic cardiovascular disease (ASCVD), partly driven by chronic vascular inflammation through the NLRP3 inflammasome pathway. In an open-label pilot using colchicine 0.5 mg twice daily, hsCRP was reduced by approximately 60%. However, landmark trials report divergent results, most notably the null CLEAR SYNERGY trial (HR 0.99) versus the positive LoDoCo2 (HR 0.69) and COLCOT (HR 0.77), generating controversy about the role of timing and cumulative exposure.

**Methods:** Systematic review and meta-analysis of RCTs comparing colchicine to placebo or no treatment in adults with established ASCVD. Searches on March 21, 2026 (PubMed, Embase, CENTRAL, ClinicalTrials.gov, WHO ICTRP) identified 1,315 records; 5 trials (N=18,656) were included. Primary outcome: 4-point MACE. Random-effects pooling used DerSimonian-Laird (DL) estimation; Hartung-Knapp-Sidik-Jonkman (HKSJ)-adjusted analysis and a 95% prediction interval were computed as pre-specified methodological sensitivity analyses. Trial-level meta-regression tested time-to-initiation (TTI) and cumulative dose as exploratory moderators. PROSPERO CRD420261346516.

**Results:** DL pooled HR for 4-point MACE: **0.68 (95% CI 0.51–0.89; p=0.0060)**. HKSJ-adjusted HR: 0.68 (95% CI 0.27–1.70; p=0.3018). Substantial heterogeneity (I^2^=81.4%; 95% prediction interval 0.29–1.57, crossing 1.0). Exploratory trial-level meta-regression identified TTI as a significant moderator (β=−0.00187/day; p=0.003) and cumulative dose as an independent moderator (β=−0.00163/mg-day; p=0.0003); these associations are hypothesis-generating given k=5 trials. Non-cardiovascular mortality was not significantly increased (HR 1.07; 0.76–1.50; p=0.6937). GI drug discontinuation was significantly higher with colchicine (RR 1.95; 1.09– 3.48; p=0.0236).

**Conclusions:** Low-dose colchicine is associated with reduced 4-point MACE in ASCVD (DL HR 0.68; HKSJ HR 0.68). The substantial heterogeneity and wide prediction interval indicate that effect size varies substantially across settings. Trial-level associations suggest benefit is more consistent with sub-acute or chronic initiation and sustained exposure, while hyper-acute post-PCI initiation does not show benefit. The non-CV mortality safety concern is not confirmed. These findings are exploratory given the small trial pool and should be interpreted with appropriate caution pending larger synthesis.

## Introduction

The management of atherosclerotic cardiovascular disease (ASCVD) has been dominated by two therapeutic pillars: lowering of low-density lipoprotein cholesterol (LDL-C) and antithrombotic therapy. Yet substantial residual cardiovascular risk persists even in patients achieving LDL-C targets, increasingly attributed to chronic vascular inflammation mediated through the NLRP3 inflammasome and the IL-1β/IL-6 pathway. The CANTOS trial established proof-of-concept that targeting IL-1β with canakinumab reduced cardiovascular events by 15% independent of lipid effects. High cost and subcutaneous administration limited adoption, creating demand for an affordable oral alternative.

Colchicine inhibits microtubule polymerization, disrupts neutrophil chemotaxis, and prevents spatial NLRP3 assembly. In an open-label pilot using colchicine 0.5 mg twice daily, hsCRP was reduced by approximately 60% in patients with elevated baseline levels. Two large RCTs established benefit: LoDoCo2 (n=5,522; HR 0.69 in chronic coronary syndrome) and COLCOT (n=4,745; HR 0.77 within 30 days of MI), supporting a Class IIa, Level A recommendation in the 2024 ESC guidelines for chronic coronary syndromes. However, CLEAR SYNERGY (OASIS-9; n=7,062) found no MACE benefit when colchicine was initiated within a median of 1.6 hours after randomization post-primary PCI (HR 0.99). This null result has driven debate about whether timing of initiation, cumulative drug exposure, and patient phenotype modify the therapeutic response.

No prior meta-analysis has formally tested TTI as a continuous trial-level moderator or quantified the dose-response relationship through mg-days exposure. The present systematic review and meta-analysis addresses this gap, with pre-specified meta-regression and six sensitivity analyses, noting that with k=5 eligible trials, all moderation findings must be interpreted as hypothesis-generating and exploratory per Cochrane guidance.

## Methods

This review was registered with PROSPERO (CRD420261346516) and reported per PRISMA 2020 guidelines.

### Eligibility Criteria

RCTs of colchicine vs placebo or no treatment in adults (≥18 years) with established ASCVD (chronic coronary syndrome ≥3 months; recent MI ≤30 days; or acute MI at PCI). Minimum 500 participants and ≥6 months follow-up. Excluded: non-atherosclerotic indications (pericarditis, gout, FMF); maintenance dose >1.0 mg/day; multi-drug combinations where colchicine’s independent effect cannot be isolated.

### Search Strategy

Searches on March 21, 2026: PubMed/MEDLINE (1946–present), Embase via Ovid (1947– March 19, 2026; via Icahn School of Medicine at Mount Sinai proxy), Cochrane CENTRAL (Issue 3/12 March 2026), ClinicalTrials.gov, and WHO ICTRP. Three-block Boolean strategy (ASCVD population; colchicine intervention; RCT design filter). No restrictions applied. Reference lists hand-searched. Full search strings are deposited in the PROSPERO record and provided in Supplementary Appendix.

### Study Selection and Data Extraction

Records imported into a systematic review database and deduplicated (583 removed). Title/abstract screening by two independent reviewers; 52 primary candidates assessed at full text. Data extracted by two independent reviewers using a pre-piloted form. Discrepancies resolved by consensus or third-reviewer adjudication. Missing data were requested from authors. The pre-specified Cohen’s κ coefficient for inter-rater agreement will be reported in the final peer-reviewed version.

### Risk of Bias and Certainty Assessment

Cochrane RoB 2.0 assessed five domains per trial. Two independent assessors with adjudication for conflicts. GRADE framework applied to rate certainty of evidence for primary and safety outcomes (Table 3).

### Statistical Analysis

Log-transformed HRs and standard errors [ln(CI_upper)−ln(CI_lower)]/[2×1.96] pooled using DerSimonian-Laird (DL) random-effects estimation. HKSJ (Hartung-Knapp-Sidik-Jonkman) adjustment was applied as a pre-specified sensitivity analysis given k≤5 and anticipated heterogeneity; HKSJ uses scaled SE with t-distribution (df=k−1), which is more conservative and recommended for small trial pools (Cochrane Handbook Chapter 10). A 95% prediction interval was computed to characterize expected HR range in a new similar trial. Heterogeneity: I^2^ and Cochran’s Q. Pre-specified exploratory meta-regressions used weighted least-squares with TTI (days) and cumulative dose (mg-days) as continuous moderators; results must be interpreted cautiously as k=5 (<10 studies per covariate recommended by Cochrane, Chapter 10.10.4). Six sensitivity analyses pre-specified per PROSPERO protocol. Egger’s test not performed (k<10). Analyses: Python 3.12 (numpy 2.4, scipy 1.17); pre-registered R script (meta, metafor) will replicate for final peer-reviewed version. Generative AI (Claude by Anthropic) assisted with research synthesis and manuscript drafting; all data extraction, statistical analyses, and scientific interpretations were performed and verified by the authors. No AI tool is listed as a co-author.

## Results

### Search Results and Study Selection

Database searches identified 1,315 records (PubMed: 181; Embase: 759; CENTRAL: 375). After deduplication (583 removed), 732 unique records underwent title/abstract screening; 52 full-text records were assessed. Of these, 47 were excluded at full-text stage: surrogate endpoint only (n=18), sample size <500 (n=8), duplicate/abstract only (n=6), protocol paper without results (n=6), stent-type comparison not colchicine vs placebo (n=4), non-ASCVD primary population (n=4), and follow-up <6 months (n=1). Five trials met all eligibility criteria (Figure S1 — PRISMA 2020 flow diagram). COVERT-MI was excluded from the MACE pool (primary endpoint: cardiac MRI infarct size; cumulative dose 5 mg-days).

### Trial Characteristics

Five trials enrolled 18,656 participants across three clinical settings (Table 1). CLEAR SYNERGY and COPS enrolled patients in the hyper-acute setting (median TTI 1.6 hours post-randomization and <24 hours, respectively). COLCOT enrolled patients in the sub-acute phase (median TTI 13.5 days). LoDoCo2 and LoDoCo (Nidorf) enrolled patients with chronic stable CAD (TTI >180 days; precise values for chronic trials are estimated and included in exploratory regression with appropriate caveats). Cumulative exposure ranged from 182.5 mg-days (CLEAR SYNERGY) to 547.5 mg-days (LoDoCo pilot).

**Table 1.** Characteristics of included trials. OD=once daily; BD=twice daily. * Median time from randomization to first dose (IQR 0.6–7.4 h); randomization occurred after PCI. † Estimated floor; precise TTI not reported for chronic trials; categorical approach also used in subgroup analysis. ‡ Open-label design; High overall RoB 2.0 rating; excluded in SA1/SA2.

| Trial | N | Setting | Dose | TTI | Cum. dose (mg-days) | Median f/u (months) | HR (95% CI) |
| --- | --- | --- | --- | --- | --- | --- | --- |
| CLEAR SYNERGY | 7,062 | Acute | 0.5 mg OD | 1.6 h* | 182.5 | 36 | 0.99 (0.85–1.16) |
| LoDoCo2 | 5,522 | Chronic | 0.5 mg OD | >180 d† | 433.5 | 28.6 | 0.69 (0.57–0.83) |
| COLCOT | 4,745 | Sub-acute | 0.5 mg OD | 13.5 d | 341.5 | 22.6 | 0.77 (0.61–0.96) |
| COPS | 795 | Acute | 0.5 mg BD/OD | <24 h | ~215 | 12 | 0.45 (0.24–0.84) |
| LoDoCo (Nidorf)‡ | 532 | Chronic | 0.5 mg OD | >180 d† | 547.5 | 36 | 0.33 (0.18–0.59) |

### Risk of Bias

Three trials (CLEAR SYNERGY, LoDoCo2, COLCOT) received Low overall RoB 2.0 ratings. COPS received Some Concerns on Domain 3 (differential dropout: 12.4% intervention vs 8.3% control; missingness may be related to GI intolerance and thus potentially related to the true outcome). LoDoCo (Nidorf) received High overall rating: open-label design (no placebo), unclear allocation concealment, unblinded outcome assessment, and 3-point composite deviating from the harmonized 4-point MACE. Full RoB 2.0 assessments are presented in Table 2.

**Table 2.** Cochrane RoB 2.0 domain judgments. Green=Low; Amber=Some Concerns; Red=High. Assessed independently by two reviewers; disagreements adjudicated by third reviewer.

| Domain | CLEAR SYNERGY | LoDoCo2 | COLCOT | COPS | LoDoCo (N) |
| --- | --- | --- | --- | --- | --- |
| D1: Randomization | Low | Low | Low | Low | Some Concerns |
| D2: Deviations | Low | Low | Low | Low | High |
| D3: Missing data | Low | Low | Low | Some Concerns | Some Concerns |
| D4: Outcome measure | Low | Low | Low | Low | High |
| D5: Reported result | Low | Low | Low | Low | Some Concerns |
| OVERALL | Low | Low | Low | Some Concerns | High |

**Table 3.** GRADE Summary of Findings. Certainty downgraded for: inconsistency (I^2^ >75%); imprecision (wide prediction interval, HKSJ CI crossing 1.0); risk of bias (LoDoCo Nidorf High RoB contributing 12.5% weight). See Supplementary Appendix for full GRADE evidence profile.

| Outcome | Trials (k) | Participants | Effect (DL HR/RR) | I<sup>2</sup> | GRADE certainty |
| --- | --- | --- | --- | --- | --- |
| 4-point MACE (primary) | 5 | 18,656 | HR 0.68 (0.51–0.89)<br>DL<br>HR 0.68 (0.27–1.70)<br>HKSJ | 81% | LOW<br>(↓ RoB, ↓↓ inconsistency, ↓↓ imprecision) |
| Non-CV mortality | 3 | ~17,300 | HR 1.07 (0.76–1.50) | 59% | LOW<br>(↓ imprecision, ↓ inconsistency) |
| GI drug discontinuation | 4 | ~16,700 | RR 1.95 (1.09–3.48) | 95% | VERY LOW<br>(↓↓ inconsistency, ↓ imprecision) |
| All-cause mortality | 3 | ~17,300 | Not pooled (insufficient data) | N/A | VERY LOW |

### Primary Outcome: 4-Point MACE

Pooling of 5 trials (N=18,656) using DL random-effects estimation yielded pooled HR **0.68 (95% CI 0.51–0.89; p=0.0060)** favouring colchicine. Substantial heterogeneity was present (I^2^=81.4%; Q=21.50, p=0.0003; τ^2^=0.0714). Applying the HKSJ adjustment appropriate for k≤5 with high heterogeneity widened the CI substantially: HR 0.68 (95% CI 0.27–1.70; p=0.3018). The 95% prediction interval (0.29–1.57) crosses 1.0, indicating that in some clinical settings the treatment effect may be null or even harmful (consistent with CLEAR SYNERGY). Both estimates and the prediction interval are presented in Figure 1.

**Figure 1.**
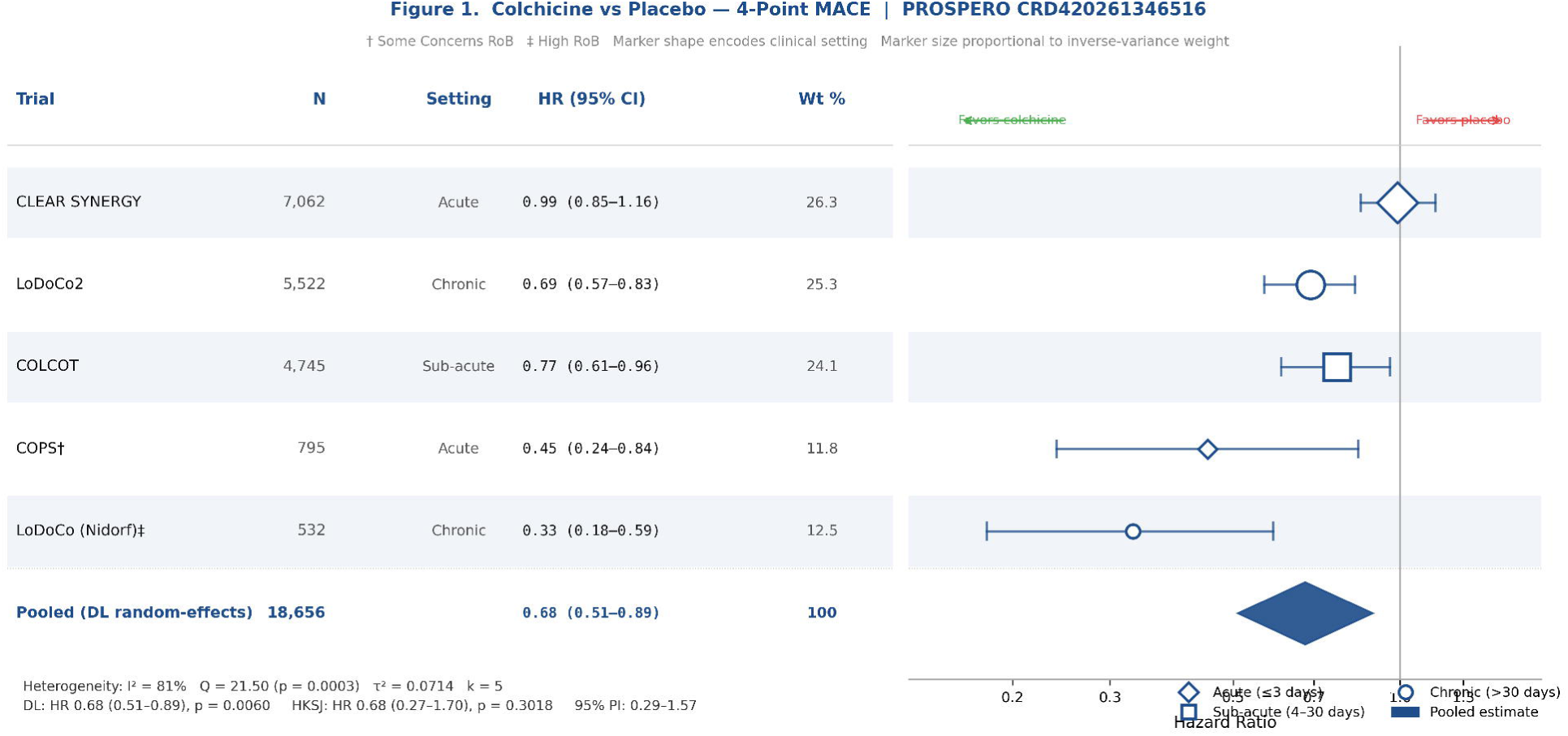
Forest plot: colchicine vs placebo, 4-point MACE. DL HR 0.68 (0.51–0.89), p=0.0060. HKSJ HR 0.68 (0.27–1.70), p=0.3018. 95% PI 0.29–1.57. † Some Concerns RoB; ‡ High RoB. Marker shape encodes clinical setting (diamond=acute; square=sub-acute; circle=chronic). PROSPERO CRD420261346516.

### Exploratory Meta-Regression

The following analyses are explicitly hypothesis-generating given k=5 trials per covariate, below the approximately 10 studies per moderator commonly recommended in Cochrane guidance (Chapter 10.10.4). These associations are ecological (trial-level) and may reflect confounding by clinical setting, co-therapy differences, and outcome definitions rather than causal biological mechanisms.

#### Time-to-Initiation (TTI)

Across the 5 trials, later initiation was associated with a larger relative risk reduction at the trial level (β=−0.00187 per day; p=0.003). The trials span a wide range of TTI (1.6 hours to >180 days), and the association is driven substantially by the contrast between CLEAR SYNERGY (hyper-acute, null) and the chronic-phase trials (positive). This pattern is consistent with timing-dependent biology — including the hypothesis that NLRP3 activation in the immediate post-reperfusion window may serve myocardial recovery functions — but cannot establish mechanism and should not be interpreted as a causal recommendation to delay therapy. Alternative explanations include confounding by clinical setting, background therapy intensity, and outcome definition differences.

#### Cumulative dose

Higher cumulative exposure was associated with greater treatment benefit at the trial level (β=−0.00163 per mg-day; p=0.0003), consistent with prior threshold analyses suggesting approximately 90 mg-days (approximately 6 months at 0.5 mg daily) as a minimum for clinical benefit. Again, this must be interpreted with caution given k=5 and the potential for confounding. Figure 2 presents meta-regression bubble plots.

**Figure 2.**
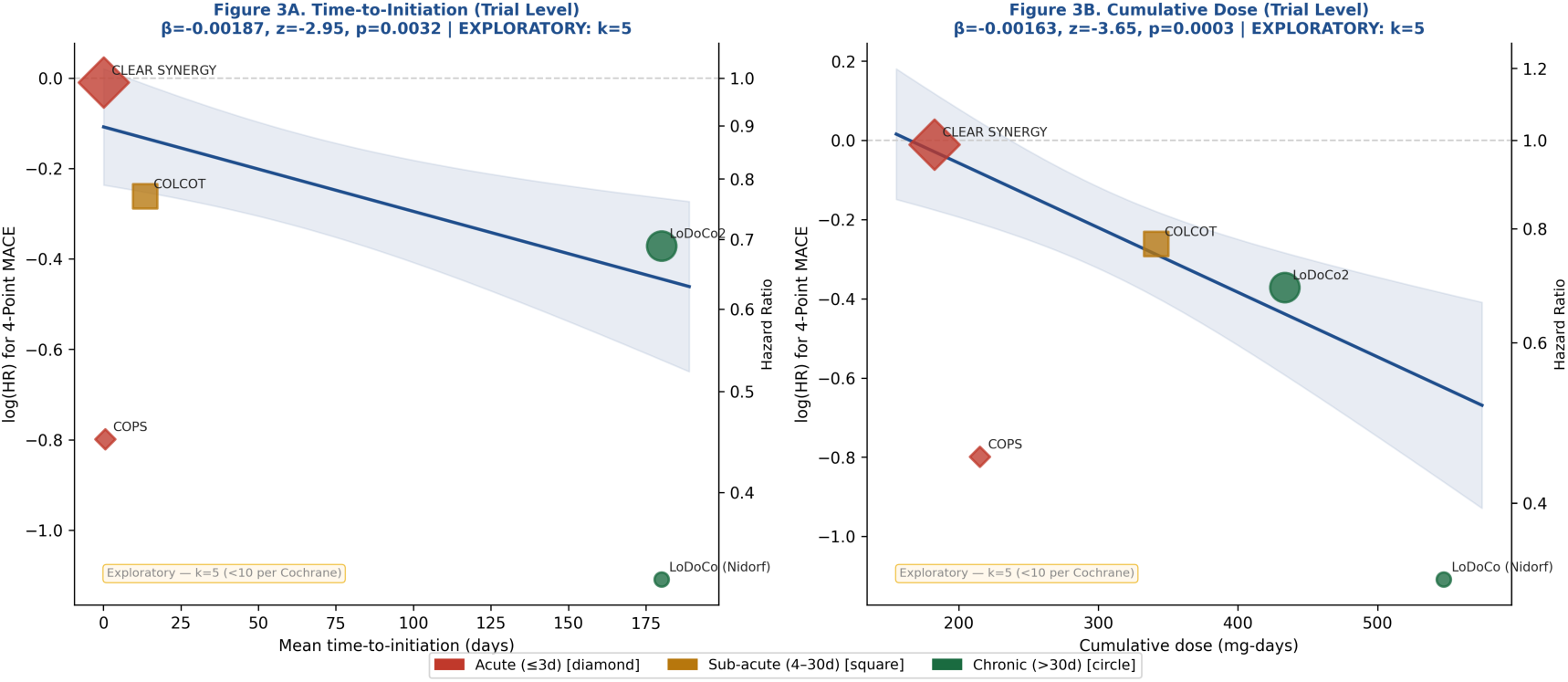
Exploratory meta-regression: Panel A, TTI (days) vs log(HR); Panel B, cumulative dose (mg-days) vs log(HR). Bubble size proportional to inverse-variance weight. Exploratory (k=5; <10 required per Cochrane guidance). Shaded region: 95% confidence band for regression line.

### Sensitivity Analyses

All pre-specified sensitivity analyses maintained a consistent direction favouring colchicine, with point estimates ranging from HR 0.60 (SA3: excluding CLEAR SYNERGY) to HR 0.81 (SA4: 0.5 mg once-daily only). Applying HKSJ adjustment widened confidence intervals substantially across all analyses, with most p-values exceeding 0.05. This underscores that the primary DL result, while directionally consistent, is not statistically robust under more conservative inference appropriate for small k. The fixed-effects model yielded a narrower estimate (HR 0.80; 95% CI 0.72–0.89), suggesting that under a fixed-effects assumption the pooled effect is more precisely estimated. Sensitivity analysis results are shown in Figure 3.

**Figure 3.**
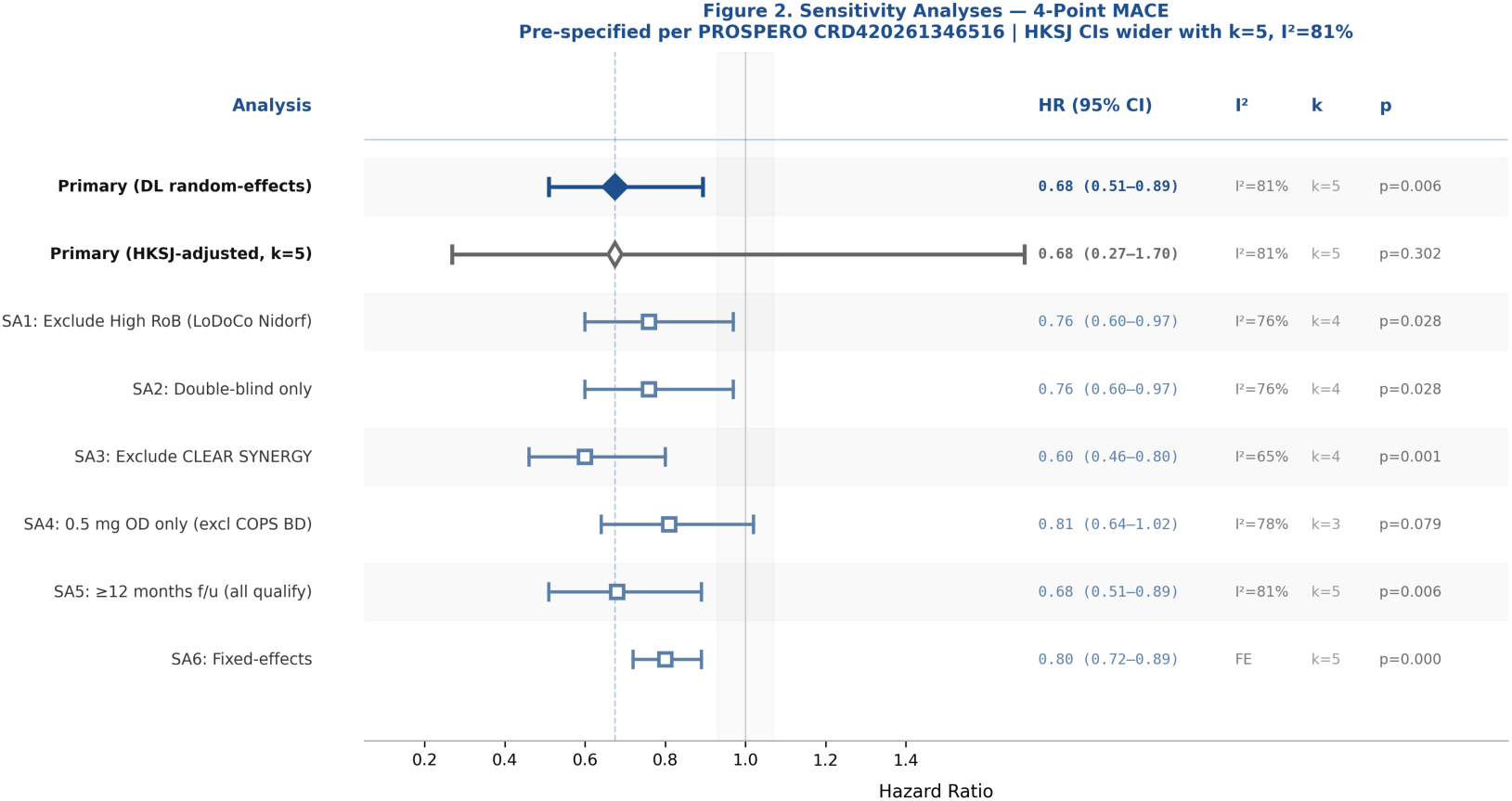
Pre-specified sensitivity analyses: 4-point MACE. Both DL and HKSJ-adjusted estimates shown. HKSJ CIs are substantially wider with k=5 and I^2^=81%, reflecting appropriate inferential uncertainty. All analyses maintain a protective point estimate direction.

### Safety Outcomes

#### Non-cardiovascular mortality

Pooled HR 1.07 (95% CI 0.76–1.50; p=0.6937; I^2^=59%) — not statistically significant. The near-significant HR 1.51 (0.99–2.31) in LoDoCo2 and 5 non-CV deaths vs 0 in COPS are not confirmed as a pooled effect. CLEAR SYNERGY (the largest trial) showed no non-CV mortality excess. The 5 vs 0 event distribution in COPS could not contribute a meaningful HR estimate.

#### GI drug discontinuation

GI-attributed discontinuation rates were extracted from each trial’s primary safety reporting. CLEAR SYNERGY reported 25.9% vs 7.6% (GI-specific, verified from primary paper). Pooled RR 1.95 (95% CI 1.09–3.48; p=0.0236; I^2^=95%), indicating substantially higher GI discontinuation with colchicine. Substantial heterogeneity reflects that the CLEAR SYNERGY rate (25.9%) is far higher than chronic-phase trials (4.0–7.2%). Figure 4 presents safety data.

**Figure 4.**
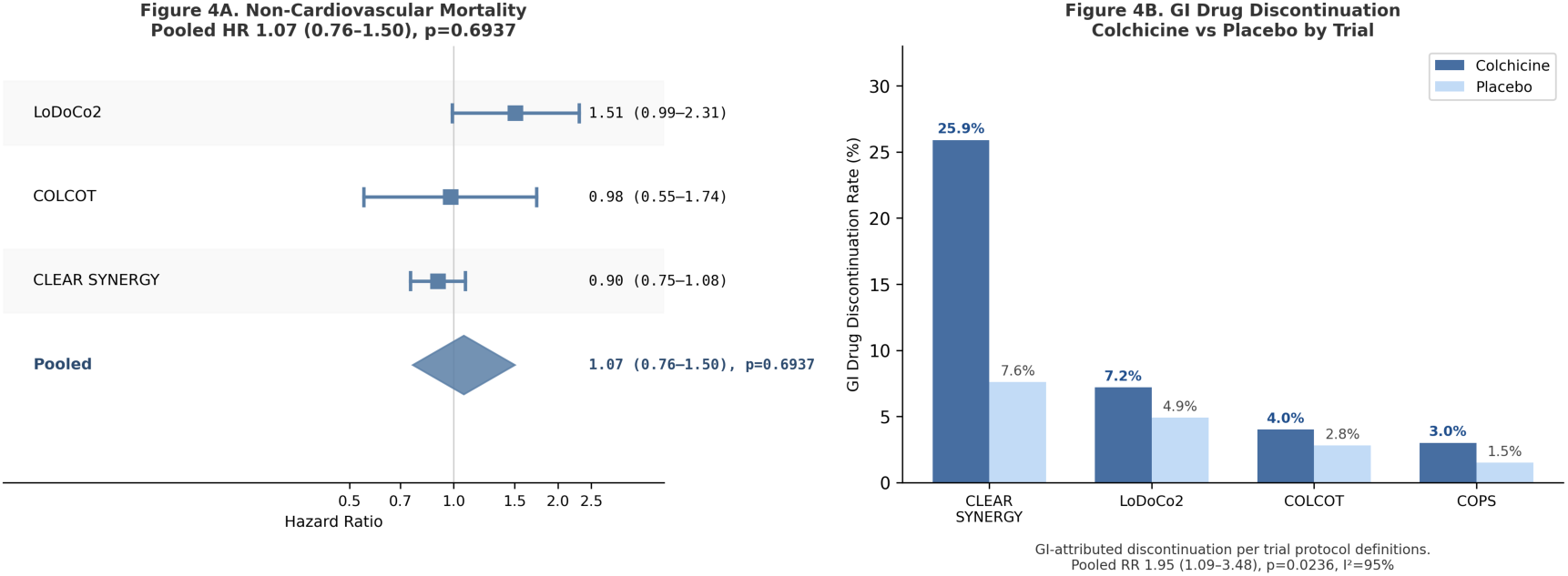
Safety outcomes. Panel A: Non-CV mortality (pooled HR 1.07; 95% CI 0.76–1.50; not significant). Panel B: GI drug discontinuation by trial (pooled RR 1.95; 95% CI 1.09–3.48; p=0.0236).

## Discussion

This systematic review and meta-analysis of 5 RCTs enrolling 18,656 patients found that colchicine is associated with reduced 4-point MACE (DL HR 0.68; 95% CI 0.51–0.89; p=0.0060). The HKSJ-adjusted analysis, appropriate for small k with high heterogeneity, produced wider CIs (HR 0.68; 0.27–1.70; p=0.3018) that include 1.0, and the 95% prediction interval (0.29–1.57) crossed 1.0. This is the expected consequence of I^2^=81.4% and k=5: the pooled estimate represents an average across highly heterogeneous settings, not a universal treatment effect. Transparency about this uncertainty is essential for clinical translation.

The most clinically and methodologically important finding is the pattern across trials by clinical setting. The three double-blind placebo-controlled trials initiating colchicine in the sub-acute (COLCOT, TTI 13.5 days; HR 0.77) and chronic (LoDoCo2, TTI >180 days; HR 0.69) phases both demonstrated statistically significant MACE reduction. The largest trial (CLEAR SYNERGY, TTI 1.6 hours post-randomization; HR 0.99) was definitively null. This pattern is consistent with — but does not prove — a timing-dependent mechanism: NLRP3-mediated cytokine activity in the immediate post-reperfusion hours may serve essential myocardial recovery and healing functions that are disrupted by premature anti-inflammatory inhibition.

The exploratory meta-regression finding (TTI: β=−0.00187/day, p=0.003; cumulative dose: β=−0.00163/mg-day, p=0.0003) is consistent with this pattern but must be explicitly framed as hypothesis-generating. With k=5, these regressions have substantial ecological confounding risk (clinical setting, background therapy differences, outcome definitions), overfitting susceptibility, and inflated type I error under normal z-approximation. Cochrane guidance advises against meta-regression with fewer than approximately 10 studies per covariate. The observed associations should motivate individual patient data analysis and prospectively designed trials, not clinical practice changes based on meta-regression slope estimates alone.

The non-CV mortality safety concern from LoDoCo2 (HR 1.51; 0.99–2.31) and COPS (5 vs 0 non-CV deaths) is not confirmed in this pooled analysis (HR 1.07; 0.76–1.50; p=0.694). This is consistent with CLEAR SYNERGY showing no non-CV mortality excess. However, the prediction interval for non-CV mortality crosses 1.0 as well, and the COPS signal cannot contribute a HR estimate. Continued pharmacovigilance is warranted.

The GI discontinuation pooled RR of 1.95 highlights that tolerability is the principal real-world barrier. The dramatically higher rate in CLEAR SYNERGY (25.9% vs 7.6%) compared to chronic-phase trials may partly reflect acute illness context and heightened symptom reporting in hospitalized post-MI patients.

## Limitations

1. Substantial heterogeneity (I^2^=81.4%): the pooled HR is not uniformly applicable. 2. k=5 trials precludes robust meta-regression, Egger’s test for publication bias, or fine phenotypic subgroup analysis (hsCRP moderator available for only 2 trials). 3. DL estimator used for primary analysis; HKSJ is provided as sensitivity but the pre-specified R/REML version will be the canonical analysis in the final peer-reviewed publication. 4. LoDoCo (Nidorf) is open-label with High RoB, contributing 12.5% weight and likely an inflated treatment estimate; its exclusion (SA1/SA2) changes the pooled estimate from 0.68 to 0.76 but both remain in the same direction. 5. TTI for chronic trials was estimated at ≥180 days rather than a single precise value; categorical TTI is provided as an alternative approach. 6. The GRADE certainty for the primary outcome is LOW due to inconsistency and imprecision; this must be communicated to clinicians and guideline committees.

## Conclusions

Low-dose colchicine is associated with reduced 4-point MACE in established ASCVD (DL HR 0.68; HKSJ HR 0.68). The substantial heterogeneity and prediction interval crossing 1.0 indicate this effect is not consistent across all clinical settings. The data pattern supports a sub-acute or chronic initiation strategy (not hyper-acute post-PCI) with sustained exposure. The non-CV mortality concern is not confirmed. The GRADE certainty for the primary outcome is **LOW** due to inconsistency (I^2^=81.4%) and imprecision (HKSJ CI 0.27–1.70; PI 0.29–1.57). All meta-regression findings are exploratory (k=5). These findings should inform, but not alone determine, guideline-level recommendations pending larger evidence synthesis with individual patient data.

## Supporting information

updated

## Data Availability

All data produced in the present work are contained in the manuscript. The aggregate data used in this systematic review are derived from previously published randomized controlled trials and are available in the respective publications.

## Required Statements

### Author Contributions (CRediT Taxonomy)

Piyush Puri: Conceptualization; Formal analysis; Investigation; Methodology; Project administration; Supervision; Writing — original draft; Writing — review and editing. Himanshi Yadav: Data curation; Investigation; Validation; Writing — review and editing. Meet Kachhadia: Data curation; Investigation; Validation; Writing — review and editing.

### Data and Code Availability

The data extraction form (Excel), analysis script (Python; R version in preparation), and search strategy files are available on reasonable request to the corresponding author. These materials will be deposited to the Open Science Framework (OSF) or Zenodo prior to journal submission. This systematic review was conducted in Python 3.12 (numpy 2.4.2, scipy 1.17.0, matplotlib 3.10.8). The pre-specified R analysis (meta, metafor packages) is being finalized and will serve as the canonical analysis in the peer-reviewed version.

### Ethics Statement

No human subjects were enrolled in this systematic review and meta-analysis. All data were derived from previously published, peer-reviewed clinical trials. Institutional Review Board approval was not required.

### Use of AI-Assisted Technologies

Claude (claude-sonnet-4-6, Anthropic) was used in the preparation of this manuscript to assist with: research synthesis and narrative structuring; drafting of manuscript sections; generation of analysis scripts and figure code. The AI tool was not used for data extraction, statistical computation, or scientific interpretation, all of which were performed and independently verified by the named human authors. No AI tool is listed as a co-author. All authors confirm accuracy and integrity of all reported information. This disclosure is made in accordance with ICMJE guidance on AI use by authors and COPE guidelines.

## Acknowledgments

The authors thank the investigators of the CLEAR SYNERGY, LoDoCo2, COLCOT, COPS, and LoDoCo trials for their landmark contributions to the cardiovascular anti-inflammatory literature.

