## Supplementary material for "Precision Anti-Inflammatory Therapy in Atherosclerosis: A Systematic Review and Meta-Analysis of Colchicine Timing and Clinical Outcomes in Patients with Atherosclerotic Cardiovascular Disease": updated

PROSPERO Registration: CRD420261346516
MS ID: MEDRXIV/2026/348968

##### TABLE OF CONTENTS

S1. Eligibility Criteria (PICO Framework)
S2. Database Search Strategies
S3. Data Extraction Template
S4. PRISMA 2020 Flow Diagram
S5. Recommended Methods Language for Manuscript

#### S1. Eligibility Criteria (PICO Framework)

This systematic review and meta-analysis followed a pre-specified protocol registered with PROSPERO (CRD420261346516). Eligibility criteria were defined a priori according to the PICO(S) framework as outlined in the table below. Independent dual screening was performed at both title/abstract and full-text stages, with discordance resolved by consensus or adjudication by a third investigator.

| PICO(S) Component | Criteria |
|---|---|
| **INCLUSION CRITERIA** | |
| **Population (P)** | Adults ≥18 years with established atherosclerotic cardiovascular disease (ASCVD), defined as one or more of the following: (1) prior myocardial infarction (MI) or acute coronary syndrome (ACS) including STEMI, NSTEMI, or unstable angina; (2) stable coronary artery disease (CAD) confirmed by coronary angiography, CT angiography, or stress testing; (3) ischemic stroke or transient ischemic attack (TIA); (4) symptomatic peripheral artery disease (PAD); (5) coronary revascularization (PCI or CABG) |
| **Intervention (I)** | Colchicine administered for cardiovascular indication, any dose and formulation (including low-dose colchicine 0.5 mg/day, standard-dose 0.6 mg twice daily, or any other dose regimen); studies must specify timing of initiation relative to index cardiovascular event (acute phase: ≤30 days post-event; chronic/maintenance: >30 days or for stable disease) |
| **Comparator (C)** | Placebo, standard of care (guideline-directed medical therapy), or active comparator (other anti-inflammatory agent). Open-label studies without a concurrent comparator group are excluded unless pre-post data allows extraction of controlled effect estimates. |
| **Outcomes (O)** | Primary: Major adverse cardiovascular events (MACE), defined as composite of cardiovascular death, non-fatal MI, and/or non-fatal stroke (3-point or 4-point MACE). Secondary: All-cause mortality; cardiovascular mortality; recurrent MI; ischemic stroke; hospitalization for unstable angina; coronary revascularization (any); C-reactive protein (CRP) or high-sensitivity CRP (hsCRP) change; interleukin-1β (IL-1β) or IL-6 levels; gastrointestinal adverse events; serious adverse events (SAE); treatment discontinuation rates. |
| **Study Design (S)** | Randomized controlled trials (RCTs) and prospective/retrospective observational studies (cohort studies, case-control studies). Minimum study size: ≥50 participants in the colchicine arm. Minimum follow-up: ≥3 months. Full-text publication required. Published from January 1, 2000 to present. All languages accepted (non-English studies translated via Google Translate with data verified by bilingual co-investigator if available). |
| **EXCLUSION CRITERIA** | |
| **Population** | Pediatric populations (<18 years); patients receiving colchicine exclusively for non-cardiovascular indications (gout, familial Mediterranean fever, pericarditis without concurrent ASCVD); patients with active inflammatory conditions as primary disease (rheumatoid arthritis, systemic lupus erythematosus, vasculitis) without confirmed ASCVD |
| **Study Design** | Case reports; case series (n<10); editorials; letters; review articles (non-systematic); conference abstracts without peer-reviewed full-text publication; studies without extractable outcome data; duplicate publications (most complete dataset retained); studies where colchicine is used as part of a multi-drug combination without an isolated effect estimate |
| **Outcomes** | Studies reporting only surrogate endpoints without any clinical cardiovascular outcome; studies reporting only pharmacokinetic or pharmacodynamic endpoints without clinical outcomes; studies with follow-up <3 months |

#### S2. Database Search Strategies

Comprehensive searches were conducted across four electronic databases and two trial registries. Searches combined controlled vocabulary (MeSH/Emtree) with free-text terms and were not restricted by language or publication status. All searches were conducted by P.P. and verified independently by a second reviewer. Search dates and hit counts are documented for each database.

##### S2.1 PubMed / MEDLINE

*Platform: PubMed (National Library of Medicine)* | *Date of Search: March 21, 2026* | *Total Records: 181*

| Line | Search String |
|---|---|
| #1 | ("Colchicine"[MeSH] OR "colchicine"[tiab] OR "colcemid"[tiab] OR "low-dose colchicine"[tiab]) |
| #2 | ("Atherosclerosis"[MeSH] OR "Coronary Artery Disease"[MeSH] OR "Myocardial Infarction"[MeSH] OR "Acute Coronary Syndrome"[MeSH] OR "Stroke"[MeSH] OR "Peripheral Arterial Disease"[MeSH]) |
| #3 | ("atherosclerosis"[tiab] OR "atherosclerotic"[tiab] OR "coronary artery disease"[tiab] OR "CAD"[tiab] OR "myocardial infarction"[tiab] OR "heart attack"[tiab] OR "STEMI"[tiab] OR "NSTEMI"[tiab] OR "acute coronary syndrome"[tiab] OR "ACS"[tiab] OR "unstable angina"[tiab] OR "ischemic stroke"[tiab] OR "peripheral artery disease"[tiab] OR "PAD"[tiab] OR "cardiovascular disease"[tiab] OR "ASCVD"[tiab]) |
| #4 | ("Cardiovascular Diseases"[MeSH] OR "Secondary Prevention"[MeSH] OR "Anti-Inflammatory Agents"[MeSH]) |
| #5 | ("MACE"[tiab] OR "major adverse cardiovascular"[tiab] OR "cardiovascular death"[tiab] OR "cardiovascular mortality"[tiab] OR "all-cause mortality"[tiab] OR "secondary prevention"[tiab] OR "clinical outcomes"[tiab] OR "recurrent MI"[tiab] OR "stroke"[tiab] OR "revascularization"[tiab]) |
| #6 | #2 OR #3 OR #4 |
| #7 | #1 AND #6 |
| #8 | #7 AND (#5) |
| #9 | #7 AND ("Randomized Controlled Trial"[pt] OR "Clinical Trial"[pt] OR "cohort study"[tiab] OR "observational study"[tiab] OR "retrospective study"[tiab] OR "prospective study"[tiab]) |
| #10 | #8 OR #9 |
| #11 | #10 AND ("2000/01/01"[PDAT] : "3000/12/31"[PDAT]) |
| FINAL | #11 [NOT limited by language] — Apply Humans filter via PubMed interface |

##### S2.2 Embase (via Ovid)

*Platform: Ovid Embase* | *Date of Search: March 21, 2026* | *Total Records: 759*

| Line | Search String |
|---|---|
| 1 | colchicine/exp OR colchicine:ab,ti OR 'low-dose colchicine':ab,ti |

| | |
|---|---|
| 2 | 'atherosclerosis'/exp OR 'coronary artery disease'/exp OR 'myocardial infarction'/exp OR 'acute coronary syndrome'/exp OR 'ischemic stroke'/exp OR 'peripheral artery disease'/exp |
| 3 | atherosclerosis:ab,ti OR 'coronary artery disease':ab,ti OR 'myocardial infarction':ab,ti OR STEMI:ab,ti OR NSTEMI:ab,ti OR 'acute coronary syndrome':ab,ti OR ACS:ab,ti OR 'unstable angina':ab,ti OR 'ischemic stroke':ab,ti OR 'peripheral artery disease':ab,ti OR ASCVD:ab,ti OR CAD:ab,ti |
| 4 | 'cardiovascular disease'/exp OR 'secondary prevention'/exp OR 'anti-inflammatory agent'/exp |
| 5 | MACE:ab,ti OR 'major adverse cardiovascular event':ab,ti OR 'cardiovascular death':ab,ti OR 'all-cause mortality':ab,ti OR 'secondary prevention':ab,ti OR 'clinical outcome':ab,ti OR 'recurrent myocardial infarction':ab,ti |
| 6 | 2 OR 3 OR 4 |
| 7 | 1 AND 6 |
| 8 | 7 AND 5 |
| 9 | 7 AND ('randomized controlled trial'/exp OR 'cohort analysis'/exp OR 'retrospective study'/exp OR 'prospective study'/exp OR randomized:ab,ti OR 'clinical trial':ab,ti OR cohort:ab,ti) |
| 10 | 8 OR 9 |
| FINAL | 10 AND [humans]/lim AND [2000-current]/lim — No language restriction |

#### S2.3 Cochrane Central Register of Controlled Trials (CENTRAL)

*Platform: Cochrane Library | Date of Search: March 21, 2026 | Total Records: 375*

| Line | Search String |
|---|---|
| #1 | MeSH descriptor: [Colchicine] explode all trees |
| #2 | colchicine:ti,ab,kw OR "low-dose colchicine":ti,ab,kw |
| #3 | #1 OR #2 |
| #4 | MeSH descriptor: [Atherosclerosis] explode all trees |
| #5 | MeSH descriptor: [Coronary Artery Disease] explode all trees |
| #6 | MeSH descriptor: [Myocardial Infarction] explode all trees |
| #7 | MeSH descriptor: [Acute Coronary Syndrome] explode all trees |
| #8 | MeSH descriptor: [Stroke] explode all trees |
| #9 | MeSH descriptor: [Peripheral Arterial Disease] explode all trees |
| #10 | atherosclerosis:ti,ab,kw OR "coronary artery disease":ti,ab,kw OR "myocardial infarction":ti,ab,kw OR STEMI:ti,ab,kw OR NSTEMI:ti,ab,kw OR "acute coronary syndrome":ti,ab,kw OR ACS:ti,ab,kw OR "ischemic stroke":ti,ab,kw OR ASCVD:ti,ab,kw OR CAD:ti,ab,kw |
| #11 | #4 OR #5 OR #6 OR #7 OR #8 OR #9 OR #10 |
| FINAL | #3 AND #11 — Publication Year: 2000-present |

#### S2.4 ClinicalTrials.gov and WHO ICTRP

*Trial Registries | Date of Search: March 21, 2026*

| Registry | Search Terms Used | Results / Notes |
|---|---|---|
| **ClinicalTrials.gov (clinicaltrials.gov)** | Condition: "cardiovascular disease" OR "atherosclerosis" OR "coronary artery disease" OR "myocardial infarction"; Intervention: "colchicine"; Status: All (including Completed, Terminated) | 112; Completed trials checked for published/unpublished results |
| **WHO ICTRP (trialsearch.who.int)** | Condition: cardiovascular disease, atherosclerosis; Intervention: colchicine | 34 (26 unique trials); Checked for international trials not captured in PubMed/Embase |

#### S2.5 Deduplication and Record Management

All records were exported and imported into Rayyan (rayyan.ai). Automated deduplication was performed followed by manual verification. Final unique records were retained for title/abstract screening.

| Source | Records (n) |
|---|---|
| PubMed/MEDLINE | **181** |
| Embase (Ovid) | **759** |
| Cochrane CENTRAL | **375** |
| ClinicalTrials.gov | **112** |
| WHO ICTRP | **34 (26 unique trials)** |
| Manual reference list screening (Nidorf 2013/LoDoCo, OPT-PEACE, D-HART2 + reference lists) | ≥3 (Nidorf 2013/LoDoCo, OPT-PEACE, D-HART2) |
| **Total before deduplication** | **1,573 (PubMed: 181; Embase: 759; CENTRAL: 375; ClinicalTrials.gov: 112; WHO ICTRP: 34; Hand search: ≥3)** |
| **Duplicates removed** | **583** |
| **Total unique records for screening** | **732** |

#### S3. Data Extraction Template

Data were extracted independently by two investigators (P.P. and Himanshi Yadav, MD) using a pre-specified structured extraction form. Discordances were resolved by consensus; a third reviewer adjudicated unresolved conflicts. Authors of included studies were contacted by email for missing or ambiguous data. The following template was used for each included study:

##### S3.A Study Identification and Design

| Variable | Extracted Value / Description |
|---|---|
| First author, year | |
| Full citation (journal, volume, pages, DOI/PMID) | |
| Country / countries | |
| Setting (single-center vs. multicenter) | |
| Study design | RCT / Prospective cohort / Retrospective cohort / Case-control / Other |
| Randomization method (if RCT) | |
| Blinding (if RCT): open-label / single-blind / double-blind | |
| Sample size calculation / power analysis reported? | Yes / No |
| Study registration (ClinicalTrials.gov ID) | |
| Funding source | Industry / Non-industry / Mixed / Not reported |
| Conflicts of interest declared? | Yes / No |
| Follow-up duration (median or mean, with IQR or SD) | |
| Study period (enrollment dates) | |

##### S3.B Population Characteristics

| Variable | Colchicine Arm | Control Arm |
|---|---|---|
| Total N enrolled | | |
| Age (mean ± SD or median [IQR]) | | |
| Male sex, n (%) | | |
| Index diagnosis (ACS / Stable CAD / Stroke / PAD / Other) | | |
| ACS type (STEMI / NSTEMI / UA) if applicable | | |
| Prior MI, n (%) | | |

| |
|---|
| Prior PCI, n (%) |
| Prior CABG, n (%) |
| Hypertension, n (%) |
| Diabetes mellitus, n (%) |
| Dyslipidemia / hypercholesterolemia, n (%) |
| Current smoker, n (%) |
| eGFR (mean ± SD, mL/min/1.73m²) |
| Baseline hsCRP (mean ± SD or median [IQR], mg/L) |
| Baseline LDL-C (mean ± SD, mmol/L or mg/dL) |
| Background statin therapy, n (%) |
| Background antiplatelet therapy (aspirin/P2Y12), n (%) |
| Background ACE inhibitor / ARB, n (%) |
| Background beta-blocker, n (%) |
| LVEF (mean ± SD, %) |

#### S3.C Intervention Details

| Variable | Details |
|---|---|
| Colchicine dose (mg/day) | |
| Dosing frequency (once daily / twice daily / other) | |
| Timing of initiation relative to index event (days) | Acute (≤30 days) / Chronic (>30 days) / Mixed |
| Duration of colchicine therapy | |
| Loading dose used? If yes, specify | Yes / No; Dose: ___ |
| Route of administration | Oral / IV / Other |
| Adherence reported? If yes, rate (%) | |
| Comparator arm description | Placebo / Standard of care / Active comparator — specify |
| Co-interventions (any anti-inflammatory added to both arms?) | |

#### S3.D Clinical Outcomes

| Outcome | Colchicine n/N (%) | Control n/N (%) | Effect Estimate (HR/OR/RR) | 95% CI / p-value |
|---|---|---|---|---|
| MACE (composite — specify definition) | | | | |
| Cardiovascular death | | | | |
| All-cause mortality | | | | |
| Non-fatal MI | | | | |
| Non-fatal ischemic stroke | | | | |
| Hospitalization for unstable angina | | | | |
| Coronary revascularization (any) | | | | |
| Emergency revascularization | | | | |
| Heart failure hospitalization | | | | |
| New-onset atrial fibrillation | | | | |
| Transient ischemic attack (TIA) | | | | |

#### S3.E Inflammatory Biomarker Outcomes

| Biomarker | Colchicine (post-tx) | Control (post-tx) | Mean Difference / % Change | 95% CI / p-value |
|---|---|---|---|---|
| hsCRP (mg/L) | | | | |
| CRP (mg/L) | | | | |
| IL-1β (pg/mL) | | | | |
| IL-6 (pg/mL) | | | | |
| IL-18 | | | | |
| Neutrophil count (×10⁹/L) | | | | |
| Leukocyte count (×10⁹/L) | | | | |

#### S3.F Safety and Adverse Events

| Adverse Event | Colchicine n/N (%) | Control n/N (%) | OR / RR | 95% CI / p-value |
|---|---|---|---|---|
| Any gastrointestinal AE | | | | |
| Diarrhea | | | | |
| Nausea / vomiting | | | | |
| Abdominal pain | | | | |

| |
|---|
| Serious adverse events (SAE) |
| Drug discontinuation (any reason) |
| Drug discontinuation (AE-related) |
| Myopathy / elevated CK |
| Neutropenia / cytopenias |
| Hepatotoxicity |
| Death attributed to drug |

##### S3.G  Subgroup Data (if reported)

| Subgroup Variable | Effect Estimate and 95% CI |
|---|---|
| Timing: acute vs. chronic colchicine | |
| ACS vs. stable CAD | |
| Dose: low (0.5 mg/day) vs. standard | |
| Age: <65 vs. ≥65 years | |
| Sex: male vs. female | |
| Diabetes: yes vs. no | |
| CRP: elevated vs. normal baseline | |
| Background statin: yes vs. no | |
| Follow-up <1 year vs. ≥1 year | |

#### S4. PRISMA 2020 Flow Diagram

The following table represents the PRISMA 2020 flow of study selection. A formal PRISMA flow figure is provided as a separate figure file (Figure S1). Green-highlighted cells indicate values confirmed from completed database searches. Remaining [n] values are to be populated upon completion of title/abstract and full-text screening.

> *Green cells = confirmed values from database searches completed March 21, 2026. Remaining [n] cells = to be filled upon completion of screening in Rayyan.*

| PRISMA 2020 Node | Count (n) |
|---|---|
| **IDENTIFICATION** | |
| Records from PubMed/MEDLINE | **181** |
| Records from Embase (Ovid) | **759** |
| Records from Cochrane CENTRAL | **375** |
| Records from ClinicalTrials.gov | **112** |
| Records from WHO ICTRP | **34 (26 unique trials)** |
| Records from manual reference screening (Nidorf 2013/LoDoCo, OPT-PEACE, D-HART2) | ≥3 (Nidorf 2013/LoDoCo, OPT-PEACE, D-HART2) |
| **Total records identified** | **1,573 (PubMed: 181; Embase: 759; CENTRAL: 375; ClinicalTrials.gov: 112; WHO ICTRP: 34; Hand search: ≥3)** |
| **Duplicate records removed** | **583** |
| **Total unique records for screening** | **732** |
| **SCREENING — TITLE / ABSTRACT** | |
| Records screened (title/abstract) | **732** |
| Records excluded (title/abstract) | **680** |
| Records advanced to full-text review | **52** |
| **SCREENING — FULL TEXT** | |
| Full-text articles assessed for eligibility | **52** |
| Full-text articles excluded — surrogate endpoint only (no clinical MACE) | **18** |
| Full-text articles excluded — sample size <500 participants | **8** |
| Full-text articles excluded — duplicate publication / abstract only | **6** |
| Full-text articles excluded — protocol paper without results | **6** |
| Full-text articles excluded — stent-type comparison (not colchicine vs placebo) | **4** |
| Full-text articles excluded — non-ASCVD primary population | **4** |
| Full-text articles excluded — follow-up <6 months | **1** |

| | |
|---|---|
| **Total full-text articles excluded** | **47** |
| **INCLUDED** | |
| **Studies included in qualitative synthesis (systematic review)** | **5** |
| Studies included in quantitative synthesis (meta-analysis) — primary outcome (4-point MACE) | **5** |
| Studies included in meta-analysis — cardiovascular / non-CV mortality | **3** |
| Studies included in meta-analysis — all-cause mortality | **3** |
| Studies included in meta-analysis — recurrent MI / stroke | **5** |
| Studies included in meta-analysis — biomarkers (hsCRP) — available in 2 trials | **2** |
| Studies included in meta-analysis — safety (GI drug discontinuation) | **4** |
| **Total participants across all included studies** | **18,656** |

*Note: The formal PRISMA 2020 flow figure (Figure S1) should be generated using the PRISMA flow diagram generator at https://www.prisma-statement.org/prismastatement/flowdiagram.aspx. Dual independent screening was performed at both stages; inter-rater reliability (Cohen's kappa) must be reported in the Methods section.*

### S5. Recommended Methods Language for Manuscript

#### Search Strategy (for Methods section)

> *We systematically searched PubMed/MEDLINE, Embase (Ovid), and the Cochrane Central Register of Controlled Trials (CENTRAL) from January 1, 2000 to March 21, 2026. Search strategies combined controlled vocabulary (MeSH/Emtree) with free-text keywords covering the population (atherosclerotic cardiovascular disease), intervention (colchicine), and clinical outcomes; full search strings are presented in Supplementary Appendix S2. We additionally searched ClinicalTrials.gov and the WHO International Clinical Trials Registry Platform (ICTRP) for registered or unpublished trials, and manually reviewed reference lists of included studies and relevant systematic reviews. No language restrictions were applied. Deduplication was performed in Rayyan (rayyan.ai), followed by two-stage independent screening by two reviewers (P.P. and Himanshi Yadav, MD) at the title/abstract and full-text levels; discordances were resolved by consensus or adjudication by a third reviewer. Inter-rater agreement was calculated using Cohen's kappa.*

#### Eligibility Criteria (for Methods section)

> *We included RCTs and observational studies (prospective and retrospective cohort designs) enrolling adults (≥18 years) with established ASCVD who received colchicine at any dose. Studies were eligible if they reported at least one pre-specified clinical outcome (MACE or individual components) with a minimum follow-up of 3 months and included ≥50 participants in the colchicine arm. We excluded case reports, conference abstracts without full-text data, and studies reporting exclusively non-clinical endpoints. Full eligibility criteria are detailed in Supplementary Appendix S1.*
